# Safety and immunogenicity of ChAd63-KH vaccine in post kala azar dermal leishmaniasis patients in Sudan

**DOI:** 10.1101/2020.08.26.20180901

**Authors:** Brima M. Younis, Mohamed Osman, Eltahir A.G. Khalil, Francesco Santoro, Simone Furini, Rebecca Wiggins, Ada Keding, Monica Carraro, E.A. Musa Anas, Mujahid A.A. Abdarahaman, Laura Mandefield, Martin Bland, Toni Aebischer, Rhian Gabe, Alison M. Layton, Charles J. N. Lacey, Paul M. Kaye, Ahmed M. Musa

## Abstract

Post kala azar dermal leishmaniasis (PKDL) is a chronic, stigmatising skin condition occurring frequently after apparent clinical cure from visceral leishmaniasis. Given an urgent need for new treatments, we conducted a Phase IIa safety and immunogenicity trial of ChAd63-KH vaccine in Sudanese patients with persistent PKDL. LEISH2a (NCT02894008) was an open label three-phase clinical trial involving sixteen adult and eight adolescent patients with persistent PKDL (median duration 30 months; range 6 -180 months). Patients received a single intramuscular vaccination of 1×10^10^ viral particles (v.p.; adults only) or 7.5×10^10^ v.p. (adults and adolescents), with primary (safety) and secondary (clinical response and immunogenicity) endpoints evaluated over 42-120 days follow up. AmBisome® was provided to patients with significant remaining disease at their last visit. ChAd63-KH vaccine showed minimal adverse reactions in PKDL patients and induced potent innate and cell-mediated immune responses measured by whole blood transcriptomics and ELISpot. 7 patients (30.4%) monitored to study completion showed >90% clinical improvement and 6 (25%) showed partial improvement. A logistic regression model applied to blood transcriptomic data identified immune modules predictive of patients with >90% clinical improvement. A randomised controlled trial to determine whether these clinical responses were vaccine related and whether ChAd63-KH vaccine has clinical utility is underway.

## Introduction

The World Health Organisation recognises the leishmaniases as some of the most significant global neglected diseases associated with poverty, with over one billion people at risk of infection with one million new cases and over 20,000 deaths reported each year^1^. These diseases are caused by infection with one of several species of the protozoan parasite *Leishmania* and are transmitted by the bite of female phlebotomine sand flies. Clinically, disease may be localised to the site of sand fly bite (cutaneous leishmaniasis; CL), spread to other skin (disseminated leishmaniasis) or mucosal (mucocutaneous leishmaniasis) sites or may involve systemic organs, notably spleen, liver and bone marrow (kala-azar or visceral leishmaniasis; VL)^3^. Post kala azar dermal leishmaniasis (PKDL), a chronic skin disease characterised by nodular or macular lesions that start on the face and spread to cover the trunk and arms, may develop in up to 50% of patients previously treated for VL^4^. PKDL is thought to play an important role in sustaining the transmission of VL, especially in inter-epidemic periods ^4-8^. Although there has been considerable success in reducing the burden of VL in South Asia following the introduction of single dose liposomal amphotericin B, this drug works less well in other geographic locations, notably East Africa, which then has led to a spate of combination drug trials involving antimonials, miltefosine, paromomycin, and amphotericin B^9^. New chemical entities and immune-modulators for VL and CL are in the early stages of clinical development but remain untested in the field^9^. There are currently no effective vaccines for the prevention or treatment of any form of leishmaniasis^11-13^.

First generation *Leishmania* vaccines composed of whole killed (autoclaved) parasites often adjuvanted with Bacillus Calmette-Guérin (BCG) were not efficacious in a prophylactic setting ^14^, but have shown signs of efficacy as an adjunct to chemotherapy for PKDL ^16^ and American cutaneous leishmaniasis^17^. Second generation vaccines that have been evaluated in clinical trials to date have been recombinant poly-protein vaccines, formulated with a variety of lipid-based adjuvants primarily aimed at eliciting CD4^+^ T cell responses (reviewed in ^19-21^), but these studies have fallen short of demonstrating efficacy in either a prophylactic or therapeutic setting. *Leishmania* as an intracellular pathogen may also be targeted for immune destruction by effector mechanisms of CD8^+^ T cells (including IFNγ production and granzyme / granulysin release ^22^) and CD8^+^ T cell responses have been associated with vaccine-induced protection in animal models ^23-26^. Vaccines designed to generate CD8^+^ T cell responses require the ability of antigen delivery into the endogenous processing pathway. This is achieved either by facilitating cross-presentation (e.g. using liposomal delivery) or through endogenous protein synthesis (e.g. naked DNA or viral vectors; so called “third generation” vaccines).

We recently described a novel third generation adenovirus-vectored vaccine (ChAd63-KH). ChAd63-KH is based on a well-characterised simian adenovirus backbone (ChAd63), extensively tested in human volunteers and shown to have an excellent safety record ^27^ ChAd-vectored vaccines induce potent CD8^+^ and CD4^+^ T cell responses and antibodies in humans and are amenable to scalable manufacture at GMP. ChAd63-KH encodes two *Leishmania* antigens, kinetoplastid membrane protein-11 and hydrophilic acylated surface protein B (K; KMP-11 and H; HASPB), both with prophylactic and therapeutic vaccine efficacy when used as monovalent vaccines in pre-clinical animal models (mouse, hamster or dog) ^24,25,28^. KMP-11 is a highly conserved membrane protein expressed in promastigotes and amastigotes of all *Leishmania* examined to date and is rich in CD8^+^ T cell epitopes ^23^. HASPB is expressed by infective metacyclics and amastigotes ^29^ and has conserved N and C termini flanking polymorphic repeats. These repeats differ in copy number and arrangement across isolates of *L. donovani* ^30^ although the functional significance of this is unknown. To increase cross-isolate coverage, we designed a synthetic KH fusion gene for ChAd63-KH, engineered to reflect HASPB repeat sequence diversity ^31^. Thus, ChAd63-KH has attributes for a pan-leishmaniasis vaccine.

Results from a first-in-human trial in UK volunteers (ISRCTN07766359) indicated that single dose vaccination with ChAd63-KH was safe, minimally reactogenic and induced potent innate and cell-mediated immune responses^32^. Here, we report on the first use of this vaccine in patients with leishmaniasis. We describe a “window of opportunity” Phase IIa clinical trial demonstrating safety and immunogenicity of single dose ChAd63-KH in Sudanese patients with persistent PKDL.

## Methods

### Ethics statement

The study was approved by the Review Committees of the Institute of Endemic Diseases, University of Khartoum, the Sudan National Medicines and Poisons Board and the Department of Biology, University of York. LEISH2a was sponsored by the University of York. The study was conducted according to the principles of the current revision of the Declaration of Helsinki 2008 and ICH guidelines for GCP (CPMP/ICH/135/95) and was registered as NCT02894008 at ClinicalTrials.gov. All participants provided written informed consent before enrolment.

### Study design and participants

LEISH2a was an open label three-phase study designed to evaluate the safety (primary endpoint), clinical response and immunogenicity (secondary endpoints) of the investigational vaccine ChAd63-KH in 24 patients with persistent PKDL of greater than six months duration. Participants were patients diagnosed with persistent PKDL aged between 18-50 (adults) or 12-16 (adolescents). The diagnosis of a case of persistent PKDL was based on a typical distribution of the skin rash for a duration of six months or more, a temporal relationship treated kala-azar, a reactive serology test and exclusion of other skin condition. PKDL lesions were defined per protocol as grade 1 (scattered maculopapular or nodular lesions, mainly around the mouth), grade 2 (dense maculopapular or nodular rash covering most of the face and extending to chest, back, upper arms and legs or grade 3 (dense maculopapular or nodular rash covering most of the body, including hands and feet.)^33^. Inclusion criteria included: uncomplicated PKDL of > 6 month’s duration; availability for the duration of the study; otherwise good health as determined by medical history, physical examination, results of screening tests and the clinical judgment of a medically qualified Clinical Investigator; negative for malaria on blood smear; judged able and likely to comply with all study requirements; willing to undergo screening for HIV, Hepatitis B and Hepatitis C; for females only, willing to undergo urinary pregnancy tests on the day of screening, on the day of vaccination (prior to vaccination) and 7 and 42 days after vaccination. Exclusion criteria included: mucosal or conjunctival PKDL; treatment for PKDL within 21 days; negative for antibodies in the RK39 strip test; receipt of a live attenuated vaccine within 60 days or other vaccine within 14 days of screening; administration of immunoglobulins and/or any blood products within the three months preceding the planned administration of the vaccine candidate; history of allergic disease or reactions likely to be exacerbated by any component of the vaccine or a history of severe or multiple allergies to drugs or pharmaceutical agents; any history of severe local or general reaction to vaccination; for females only, pregnancy, less than 12 weeks postpartum, lactating or willingness/intention to become pregnant during the study and for 3 months following vaccination; seropositive for hepatitis B surface antigen (HBsAg) or Hepatitis C (antibodies to HCV); any clinically significant abnormal finding on screening biochemistry or haematology blood tests or urinalysis; any confirmed or suspected immunosuppressive or immunodeficient state, including HIV infection; asplenia; recurrent, severe infections and chronic (more than 14 days) immunosuppressant medication within the past 6 months; tuberculosis, leprosy, or malnutrition (malnutrition in adults defined as a BMI <18.5, and in adolescents (12-17yrs) as a Z score cut-off value of <-2 SD); any other significant disease, disorder or finding, increasing risk to the volunteer, likely to influence the result of the study, or the volunteer’s ability to participate.

Participants were recruited from an endemic area in Gedaref state, Sudan and all study procedures were conducted at the Professor El-Hassan’s Centre for Tropical Medicine, Dooka, Sudan. Monitoring of the study was performed under contract by ClinServ (http://www.clinserv.net).

### Vaccine and study procedures

The clinical vaccine lot (B0004) was manufactured by Advent Srl. (Pomezia, Italy) as described in detail elsewhere^32^. The vaccine is a sterile aqueous buffered solution containing ChAd63-KH at a concentration of 7.5×10^10^ viral particles (v.p.) per ml. ChAd63-KH was administered as a single dose in 1ml volume intramuscularly into the deltoid muscle. The first eight adult volunteers received 1×10^10^v.p. and the subsequent eight adult volunteers received 7.5 x10^10^v.p.. Based on a review of the clinical data, eight adolescents were vaccinated with 7.5×10^10^ v.p.. A cautious stepwise approach was taken during vaccination with the first participant at each dose being followed for 21 days before vaccination of subsequent participants in that cohort. Patients were monitored in hospital for 7 days post vaccination and thereafter as out-patients on days 21, 42, 90 and 120 post vaccination (depending on cohort). An independent data safety and monitoring board (DSMB) meeting was held at the end of each cohort to review data and provide advice to the Sponsor regarding continuation of the trial. Clinical and biochemical test abnormalities were graded according to the protocol, based on NIH guidelines. Treatment for adverse events was provided as required. The natural history of PKDL in Sudan indicates that patients with disease persistent for greater than six months are unlikely to self cure^34^. A final end point for clinical response was scheduled to be made between day 42 and day 90 (see Results) Patients with less than 75% improvement were offered standard treatment with AmBisome® (2.5mg/kg/day for 20 days), those with between 75-90% improvement were offered conservative treatment or AmBisome®, and those with greater than 90% clinical improvement were deemed to not require further treatment. Standard treatment with AmBisome® (20 days; 2.5mg/kg/day) was provided in hospital and patients were confirmed as clinically cured at the end of treatment. Some patients defaulted from scheduled visits and were evaluated and treated at unscheduled visits based on their availability. Decisions to treat and evaluation of PKDL were performed by two experienced clinicians based on a subjective assessment of overall clinical improvement in the patient’s skin condition. Photographs of PKDL lesions were independently reviewed to confirm degree of clinical improvement.

### Whole blood transcriptomic analysis

Whole blood samples (2.5 ml) were collected into PAXGene tubes immediately prior to vaccination and at 1, 3 and 7 days post vaccination. All reagents and equipment for these analyses were supplied by ThermoFisher Scientific and processes carried out per manufacturers’ protocols, unless otherwise stated. Total RNA was extracted using the PAXgene Blood RNA kit (PreAnalytiX, QIAGEN). RNA was quantified using the Qubit® 2.0 Fluorometer with the RNA HS Assay Kit. ~50 ng of total RNA was used to construct sequencing libraries with the Ion AmpliSeq™ Transcriptome Human Gene Expression Kit. Libraries were barcoded, purified with 2.5 X Agencourt AMPure XP Magnetic Beads (Beckman Coulter) and then quantified using Ion Library TaqMan Quantitation Kit on a QuantStudio 5. Libraries were diluted to a concentration of about 50 pMol and pooled in groups of 8 for sequencing on Ion PI Chips. Chips were loaded using the Ion Chef System and the IonPI Hi-Q Chef Kit. Sequencing was performed on an Ion Proton Sequencer using Ion PI Hi-Q Sequencing 200 Kit. Data will be deposited in GEO at publication.

Differential gene expression analysis was performed using DeSeq2^35^. After count data normalization, differential gene expression analysis was performed using pooled day 0 data from the three study cohorts as the baseline for all contrasts. Enrichment of blood transcription modules at each time point in the different groups was assessed with the tmod R package^36^, using as an input the lists of differentially expressed genes ranked by the p-value after multiple test correction, as computed by DeSeq2. Significance of module enrichment was assessed using the CERNO statistical test (a modification of Fisher’s combined probability test) and corrected for multiple testing using the Benjamini–Hochberg correction. In order to identify the modules with highest correlation to the clinical response, the Z-score of each module was used to train a 1-dimensional Logistic Regression model. After 100 bootstraps the modules were ranked according to the average prediction score. Data analyses were performed by python scripts using the scikit-learn python library^37^. Gene set enrichments were performed in EnrichR^38^ and pathway analysis was conducted using IPA (QIAGEN Inc., https://www.qiagenbioinformatics.com/products/ingenuity-pathway-analysis)^39^.

### Assessment of vaccine-induced immunity

Ex vivo re-stimulation of frozen PBMC to elicit vaccine-induced CD8^+^ T cell responses was performed using Multiscreen IP ELISpot plates (Millipore), human IFNγ SA-APL antibody kits (Mabtech) and BCIP-NBT-plus chromogenic substrate (Moss Inc) as previously described^32^. Peptide re-stimulation was restricted to a peptide pools corresponding to the entire KMP-11 sequence and the N terminal conserved domain of HASPB1 (pools 1 and 2^32^) due to limitations in cells obtained from patients. Responses to medium only negative controls were subtracted and responses >50 SFC / million above the pre-vaccination response were regarded as positive. Antibody responses will be reported at a later date due to inaccessibility of trial samples at the current time due to COVID-19.

### Statistical analysis

Twenty four was chosen as an appropriate sample size for an early Phase II vaccine study to evaluate safety outcomes and explore the magnitude of outcome measures. It was not formally derived to show significant differences from any comparisons. All baseline data were summarised descriptively. Continuous measures are reported as averages (n, mean, standard deviations, median, IQR, min, max) while categorical data are reported as counts and percentages. Number of local and systemic AEs per participant are presented as median, minimum, maximum and IQR. The median number of AEs per participant (separately for local and systemic events) were compared between groups using the Mann-Whitney U test (three comparisons for each type of event). Analyses were performed using Stata v16^40^ and R v3.5.3^41^. ELISpot data were evaluated for normality using the D’Agnostino and Pearson test and compared using non-parametric Kolmogorov-Smirnov test. All analysis was performed in GraphPad Prism for MacOS v8.4.1.

## Results

### Study population and vaccine safety

Thirty-nine patients were screened for eligibility between November 2016 and April 2019 (**Figure 1**) and 24 patients with the demographic and baseline biochemical and hematological characteristics shown in **Table 1** and **Table S1** were enrolled in the study. In each of the adult cohorts, there were six patients with grade 1 PKDL and two patients with grade 2 PKDL. In the adolescent cohort, there were two patients with grade 1 PKDL, five patients with grade 2 PKDL and one patient with grade 3 PKDL. Median duration of PKDL in the study population was 30 months (range 6 -180 months). In the adult high dose, adult low dose and adolescent cohorts respectively, 4 of 8 (50%), 6 of 8 (75%) and 3 of 8 (37.5%) patients had had PKDL for >12 months duration. Twenty two of all 24 patients were followed up at the scheduled D90 visit, and 5 of 8 patients in the adolescent cohort were followed up to their D120 scheduled visit (**Figure 1**).

**Figure 1.**
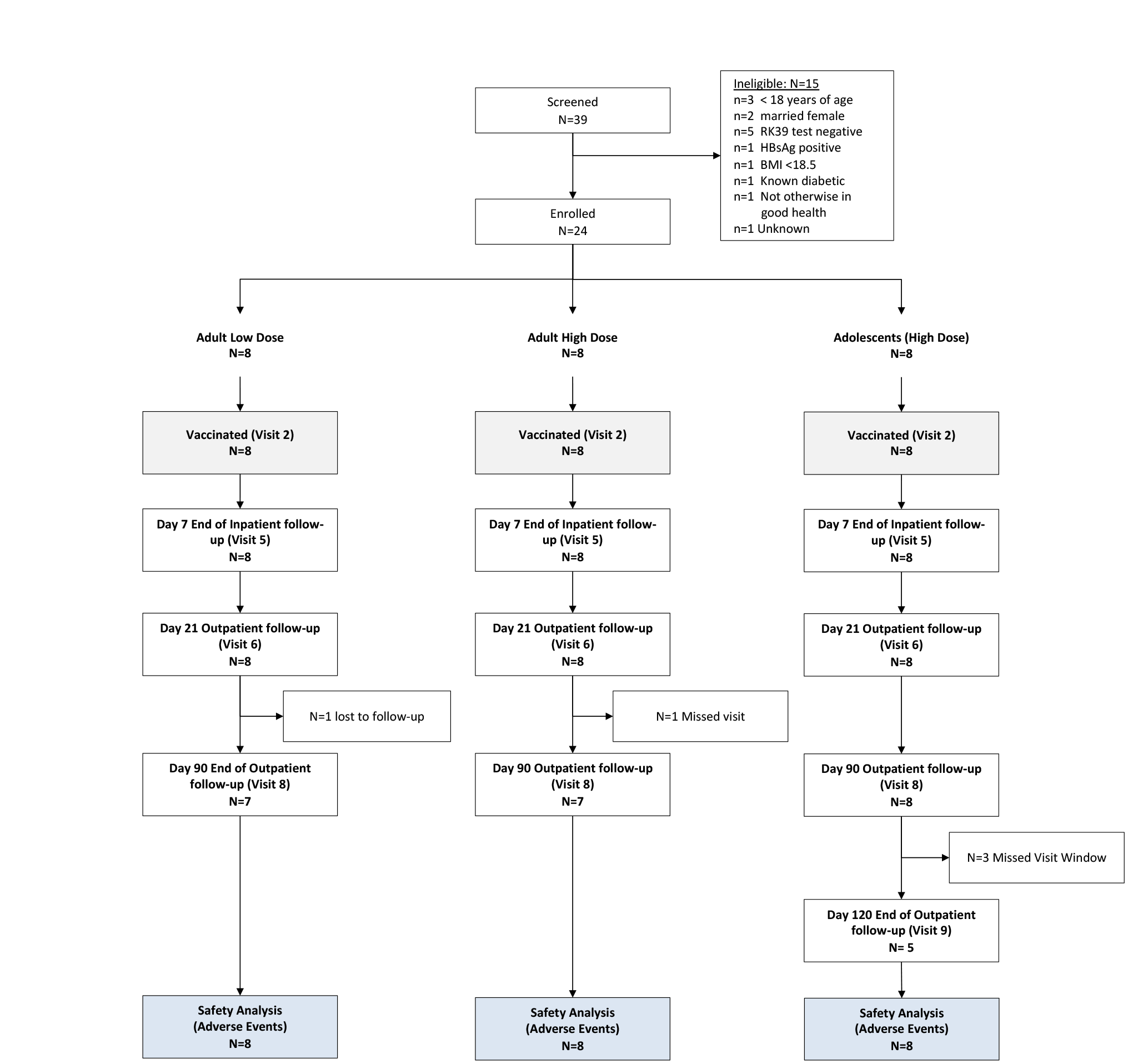
CONSORT diagram for the LEISH2a clinical trial. The CONSORT diagram reports attendance at scheduled in-patient and out-patient visits. Clinical data was also collected for some individuals at additional unscheduled visits as shown in **Figure 3**.

**Table 1.** Demographics of study population. See Table S1 (Sheet 1) for further details * Data presented as median (Q1, Q3)

|  | **Adults - Low**<br>**Dose (N=8)** | **Adults - High**<br>**Dose (N=8)** | **Adolescents**<br>**(N=8)** | **Total**<br>**(N=24)** |
| --- | --- | --- | --- | --- |
| **Sex** |  |  |  |  |
| Male | 7 (87.5%) | 8 (100.0%) | 6 (75.0%) | 21 (87.5%) |
| Female | 1 (12.5%) | 0 (0.0%) | 2 (25.0%) | 3 (12.5%) |
| **Age (years)\*** | 18.00<br>(18.00, 19.75) | 23.50<br>(19.50, 32.00) | 12.50<br>(12.00, 14.25) | 18.00<br>(14.75, 23.25) |
| **Height (m)\*** | 1.68<br>(1.63, 1.71) | 1.73<br>(1.69, 1.76) | 1.48<br>(1.37, 1.57) | 1.65<br>(1.55, 1.72) |
| **Weight (kg)\*** | 54.00<br>(51.25, 56.75) | 58.50<br>(56.25, 61.00) | 34.00<br>(30.50, 41.88) | 52.50<br>(43.62, 58.25) |
| **Duration PKDL**<br>**(months)\*** | 30<br>(6, 57) | 51<br>(18.75, 102) | 10<br>(7, 33) | 30<br>(7, 52.5) |

There were a total of 54 (8 local and 46 systemic) AEs reported during the study of which 20 (8 local and 12 systemic) were considered possibly, probably or definitely related to vaccination (**Figure 2** and **Table S1**). These included local itch (1 patient), soft swelling (2 patients) and pain (5 patients) as well as single cases of systemic malaise, pain, iron deficiency anaemia and immunological changes (leucopenia, neutropenia, thrombocytopenia and thrombocytosis). Two patients reported headache and feeling hot. Overall, 7 participants experienced at least one local and 19 experienced at least one systemic AE, with no significant differences in number of events per person between cohorts (Median adult low dose: 0 local, 2 systemic; Median adult high dose: 0 local, 1.5 systemic; Adolescent high dose: 0 local, 1.5 systemic). AEs were limited to grade 1 and 2, with no grade 3, SAEs or SUSARs reported (**Table S1**). All local and systemic AEs reported were deemed to be not serious. Medication was not required for any of the local AEs reported.

**Figure 2.**
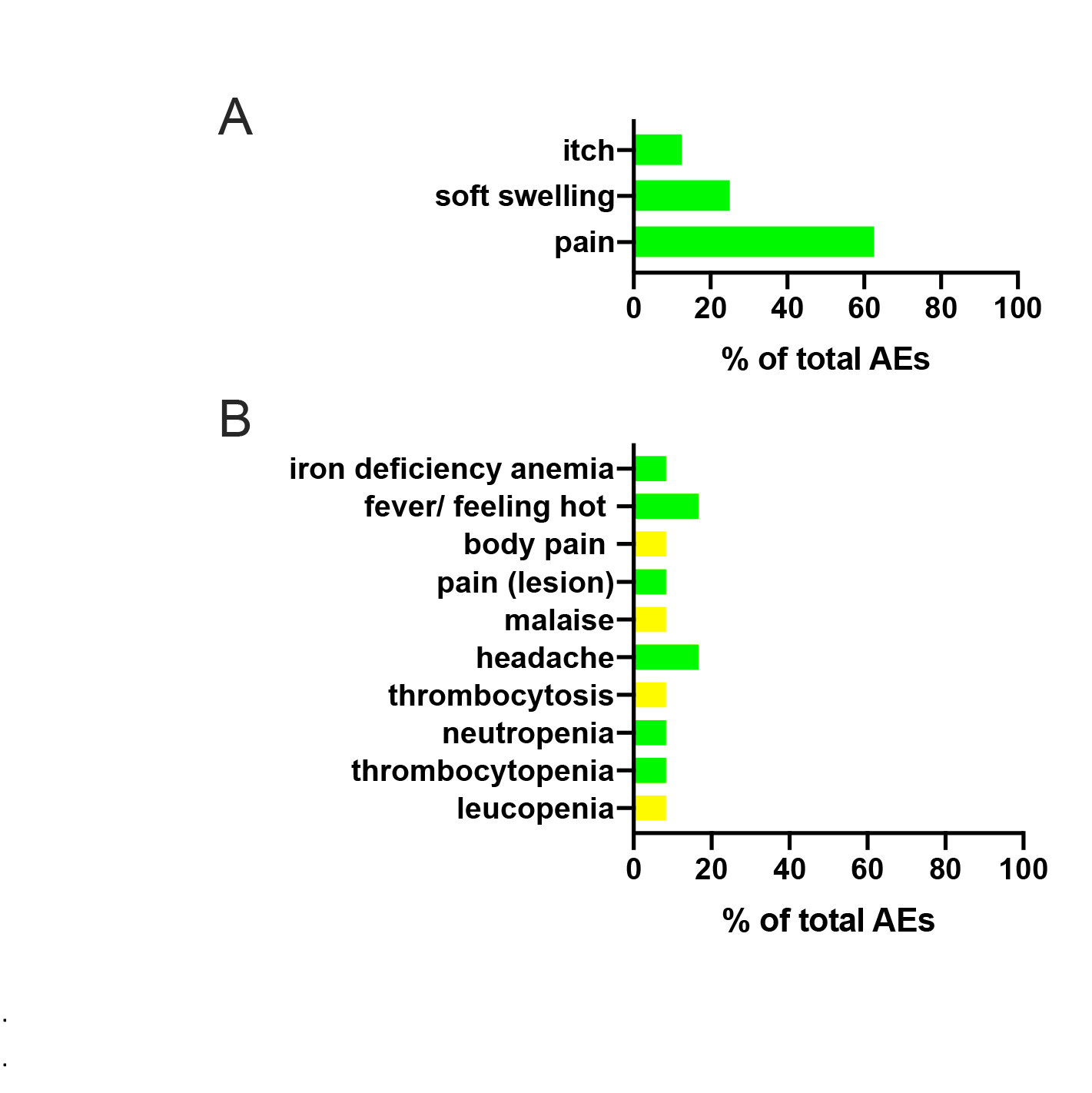
Summary of AEs reported in this study. AEs that were possibly, probably or definitely related to vaccination are shown by category as percentage of total across all three cohorts. **A**. local adverse events (n=8). **B**. systemic adverse events (n=12). Grade 1 mild, green bars. Grade 2 moderate, yellow bars.

### Clinical follow up

PKDL was subjectively assessed for each individual at each study visit and scored as percentage change in clinical disease relative to time of vaccination (**Figure 3**). Some patients were unable to attend their formal visits and/or refused treatment at that time due to harvest or schooling commitments and were examined at unscheduled visits. Based on the observation of late cure in some patients in the first two cohorts, the DSMB also approved a formal extension of the follow up period to assess cure post vaccination from 42 days to 42-90 days. Based on collective analysis of scheduled and unscheduled visits, 11/24 (45.8%) of patients had less than 25% clinical improvement and 6/24 (25%) showed clinical improvement of between 40-60% over the period of follow up. All were treated according to protocol, with the exception of one patient lost to follow up. Treatment was not required in 2 of 7 (28.6%) patients in the adult low dose cohort (both grade 1 PKDL at vaccination; **Figure 3A**), 3 of 8 (37.5%) patients in the adult high dose cohort (two grade 1 PKDL and one grade 2 PKDL at vaccination; **Figure 3B**) and 2 of 8 (25%) patients in the adolescent cohort (one grade 2 PKDL and one grade 3 PKDL at vaccination; **Figure 3C**) who reached the clinical threshold of >90% improvement in their PKDL. Overall, 7 of 23 (30.4%) patients followed to study completion resolved their PKDL lesions without the need for chemotherapy.

**Figure 3.**
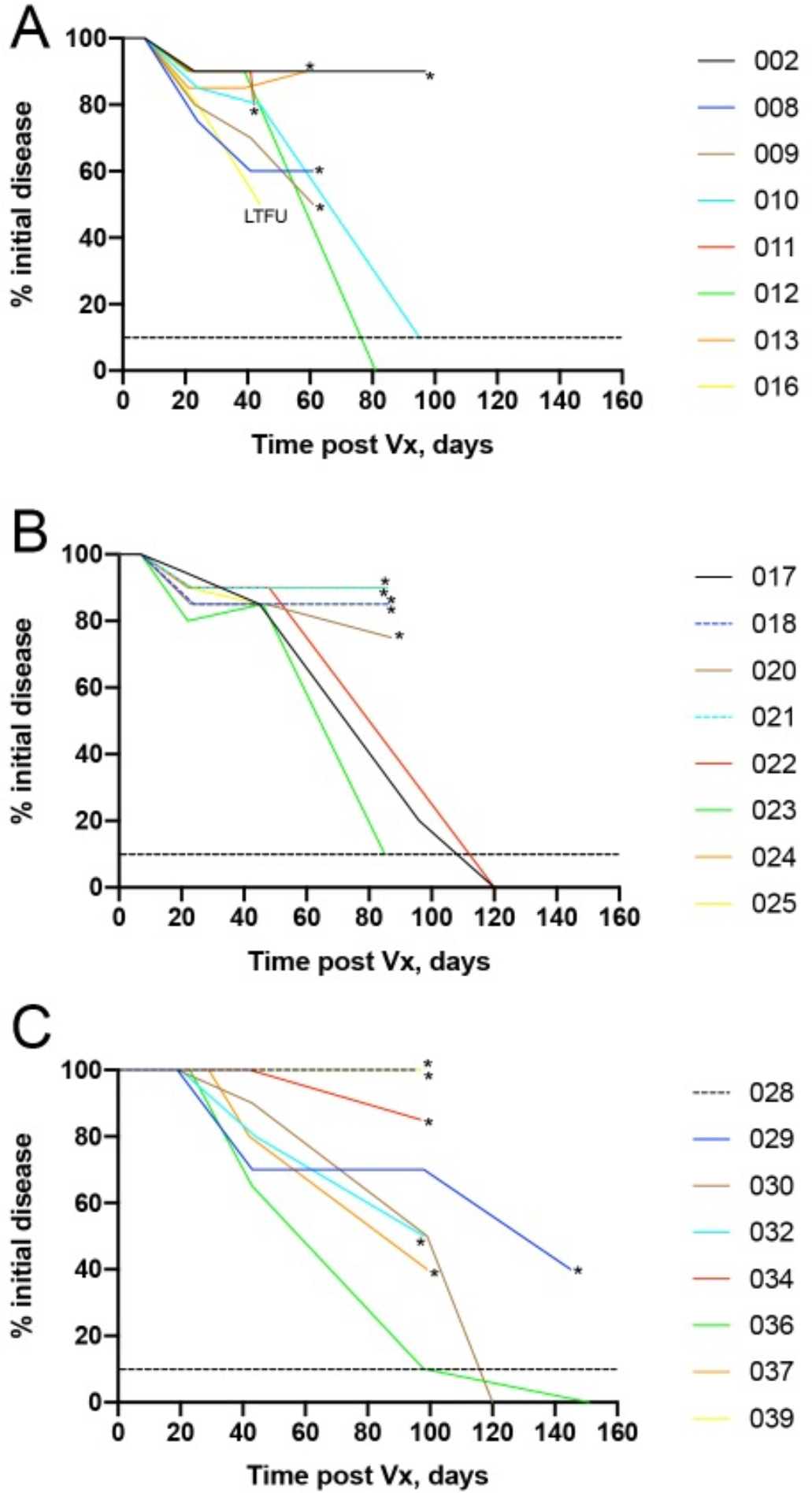
Clinical outcome for LEISH2a. Data are presented for each patient as percentage improvement of PKDL over time post vaccination, normalised to the day of vaccination. **A**. Low dose adult cohort. **B**. High dose adult cohort. **C**. High dose adolescent cohort. Asterisks indicate patient received conventional treatment with AmBisome®. LTFU: lost to follow up. Dotted line represents 90% clinical improvement.

### Whole blood transcriptome prior to and after vaccination

Immune responses in the patient cohort were assessed using whole blood transcriptomic analysis (WBTA), comparing pre-vaccination blood to blood taken at 1, 3 and 7 days post vaccination. Differentially expressed genes were identified (**Table S2**) and used to identify transcriptional modules significantly associated with vaccination (**Figure 4** and **Table S3**). These data identified three key features of the response. First, modules associated with an anti-viral signature and with dendritic cell activation showed a marked dose dependence, being minimal in patients receiving low dose vaccine. Second, there was a near equivalence of these innate responses in adults and adolescents receiving high dose vaccine. Third, modules associated with B cell responses were prominent only in adolescents. As approximately 30% of patients resolved their PKDL over the follow up period, we sought to identify potential predictors of resolution. We used a logistic regression model to identify potential predictors of patients with >90% clinical resolution, using the Z scores associated with the blood transcription modules (**Figure 4**). This analysis revealed 11 modules (Predictive modules sheet in **Table S3**), focused on monocyte and dendritic cell attributes, identified to have highest predictive value for clinical response. Among these 11 modules, two were also identified as differentially expressed: LI.M139 (lysosomal/endosomal proteins) and LI.M118.0 (enriched in monocytes) (**Table S3**).

**Figure 4.**
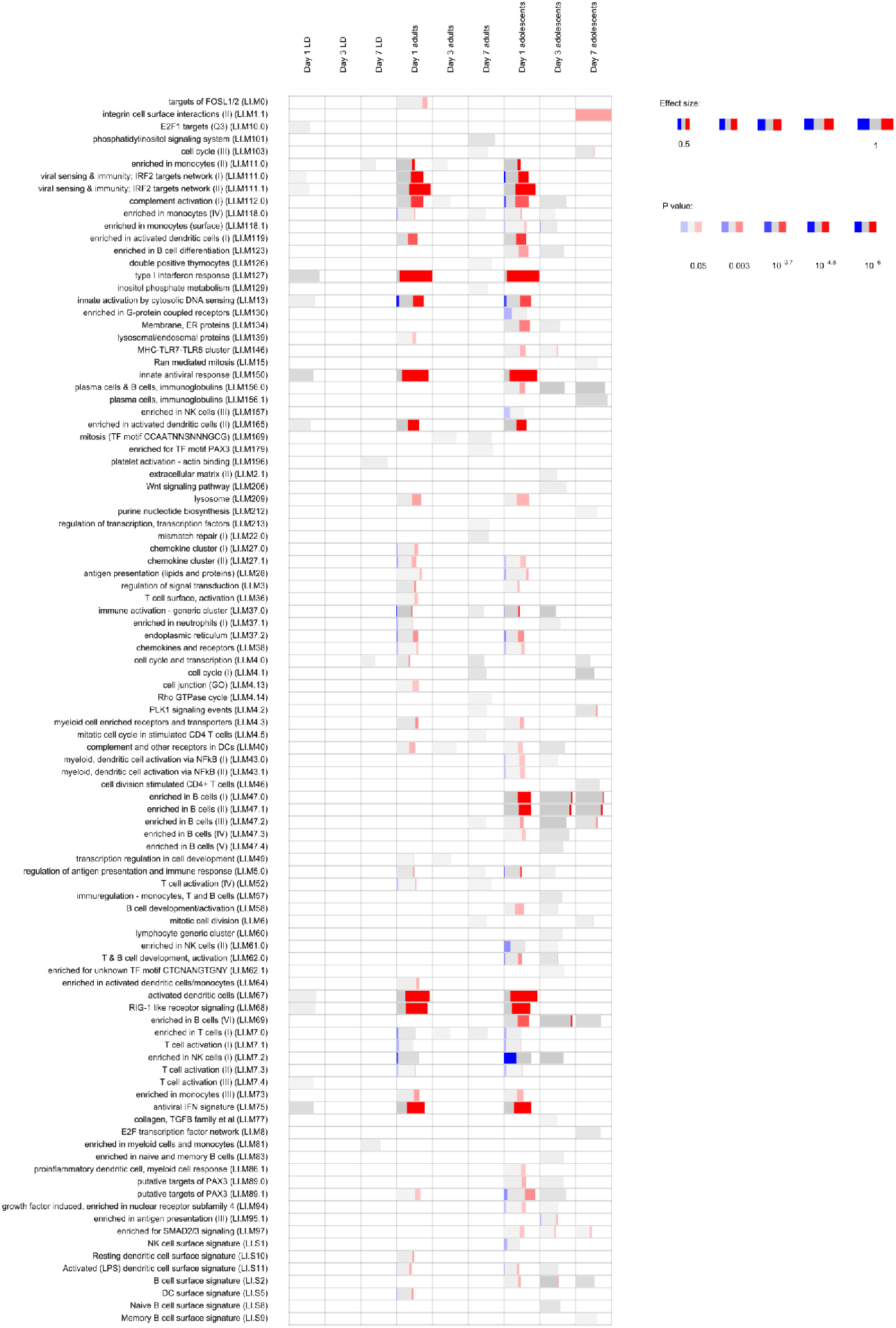
Whole blood transcriptomic analysis of patient responses to vaccination with ChAd63-KH. WBTA was conducted using the Ion AmpliSeq™ Transcriptome Human Gene Expression Kit. Each column represents a different time-point (days 1, 3, and 7) after vaccination in the three study groups (low-dose adults, high-dose adults, high-dose adolescents). Significantly enriched immune-related modules were identified applying the CERNO test on the adjusted p-value-ranked lists of genes generated by DeSeq2 (see **Table S2** for module gene lists). Modules are represented by bars in which the proportion of significantly upregulated and downregulated genes is shown in red and blue, respectively. The gray portion of the bar represents genes that are not significantly differentially regulated. The significance of module activation is proportional to the intensity of the bar, while the effect size is proportional to its width.

We also analysed differentially expressed genes identified at day 1 post vaccination with 7.5×10^10^ vp by Ingenuity Pathway Analysis and for gene set enrichment (using EnrichR). Comparative pathway analysis of differentially expressed genes in adults (374 UP; 108 DOWN) and adolescents (510 UP; 439 DOWN) showed a high degree of concordance in predicted upstream regulators, e.g. *IFNG* (z score of 9.169 vs 8.847 for adults vs. adolescents), *IFNA2* (z score of 7.834 vs. 7.925) and the transcription factor *IRF7* (z score of 7.424 vs. 7.35). In EnrichR, we similarly identified enriched GO and Reactome pathways related mainly to interferon type I and II signalling, anti-viral response, myeloid cells, phago-lysosomal functions and class I MHC mediated antigen processing and presentation (**Table S4**), in keeping with the results of the modular analysis described above. Thus, patients with PKDL appear to mount effective innate cellular responses to ChAd63-KH.

### CD8^+^ T cell response following vaccination

IFNγ production by CD8^+^ T cells was measured using ELISpot following re-stimulation with peptide pools. The frequency of CD8^+^ T cells producing IFNγ specifically in response to KMP-11 (pool 1) was not significantly different between high and low dose vaccinated adults nor between adult and adolescent cohorts (**Figure 5A**). Overall ELISpot responder frequency to this peptide pool was 75% (18/24), with peak responses in responders ranging from 86-3766 SFC/million PBMC (mean 509; 95% CI 93-926) (**Figure 5B**). Responses were comparable in frequency and magnitude to those seen previously in healthy UK volunteers (**Figure 5C**). Cells from adolescent patients were also evaluated for response to the N terminal of HASPB1 (pool 2; 5/8 responders; mean 461; 95% CI -45-967), again showing comparable results to that seen in UK volunteers (**Figure 5D**). These data indicate that PKDL patients respond with similar vigour to the ChAd63-vectored vaccine antigens as previously observed in healthy UK volunteers.

**Figure 5.**
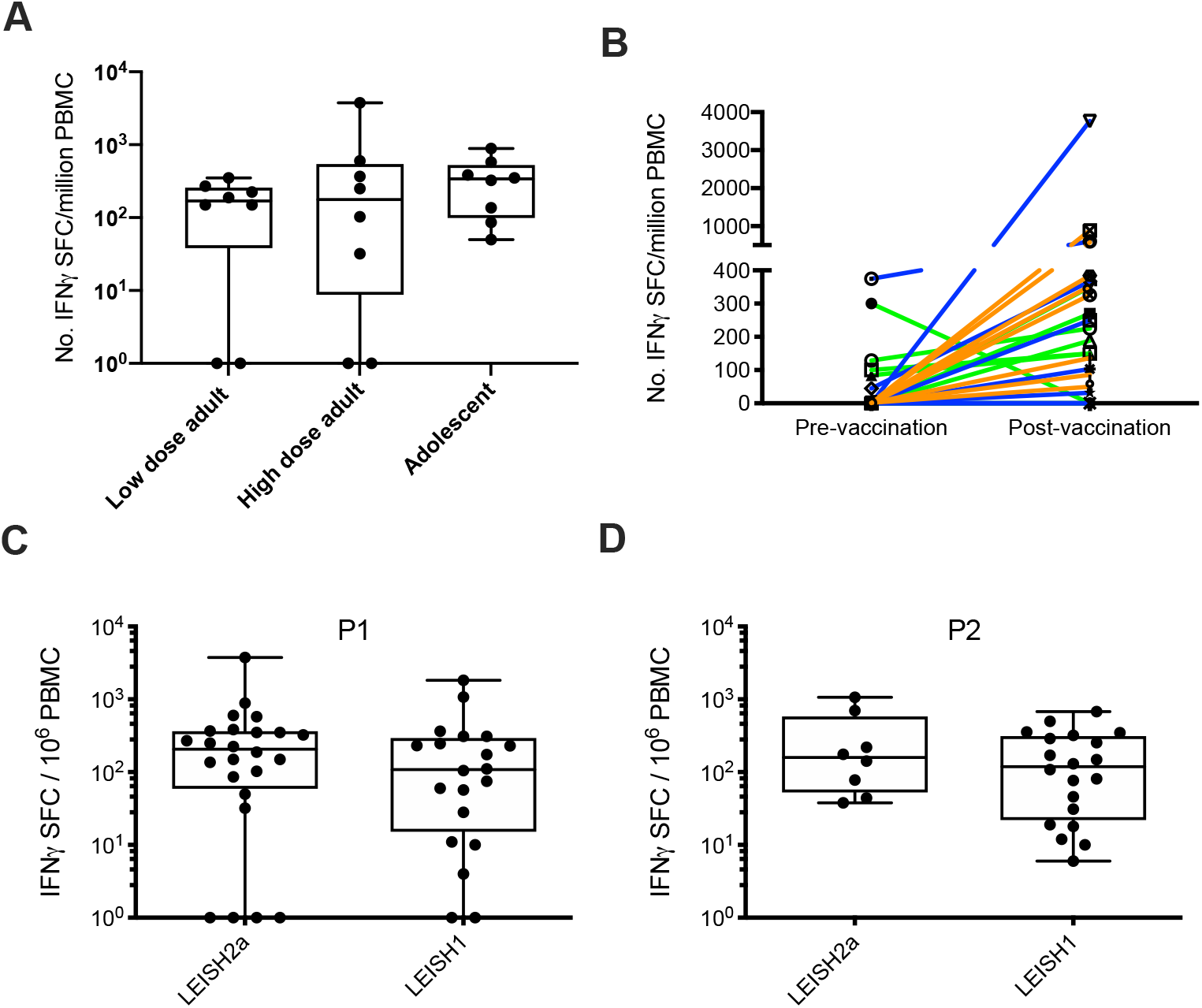
CD8^+^ T cell response to vaccination with ChAd63-KH. PBMC from patients collected from d7-d90 post vaccination were stimulated with peptide pools representing the entire KMP-11 sequence (P1) and the HASPB N terminus (P2). The number of IFNγ producing cells / million PBMC was determined by ELISpot. **A**. Peak response by cohort to P1. **B**. Pre-vaccination and peak post vaccination response per patient to P1 for low dose adult (green), high dose adult (blue) and high dose adolescents (orange); **C and D**. Comparison between patients in this trial (LEISH2a) and healthy UK volunteer responses (LEISH1) for response to P1 (C) and P2 (D). Data for LEISH1 is taken from ^32^. Box and whisker plots indicate median, 25^th^ -75^th^ quartiles, mix/max values and individual patient data points.

## Discussion

Previous studies in Sudan pioneered the use of immunotherapy with first generation vaccines (autoclaved *L. major* /Alum ^+^ BCG) in combination with sodium stibogluconate^16^, but use of a vaccine as monotherapy in PKDL patients has not been previously reported. In this Phase IIa dose escalation, age de-escalation clinical trial we have demonstrated that ChAd63-KH is both safe and immunogenic in Sudanese PKDL patients, setting the scene for further studies aimed at evaluating efficacy in a therapeutic setting.

The rationale for using vaccines for therapeutic benefit or for post exposure prophylaxis is not new and has been evaluated for a variety of chronic viral infections including HIV^43^ and HPV^44^ as well as in cancer ^45^. Whilst some chronic viral infections may subvert immunity to such an extent to pose a barrier to therapeutic vaccination^46^, recent evidence suggest that such limitations to vaccine efficacy can be overcome^47^. Evidence also suggests leishmaniasis patients may respond well to therapeutic vaccination. For example, protective immunity is readily reactivated after drug cure ^11^; virally-vectored antigen delivery generates effector CD8^+^ T cells and therapeutic benefit in rodents^31^; and there have been encouraging data from human immunochemotherapy trials in leishmaniasis patients ^16^

PKDL in Sudan has a complex natural history^5-7^. In most cases it emerges within 3-6 months of the cessation of treatment for VL, reminiscent of an immune reactivation disease^5^ although cases also occur during and even in the absence of prior VL^4^. In a study of the natural history of PKDL in 134 children younger than 14^34^, 84% of patients showed spontaneous remission of their disease with a mean (SD) of 9.7 (4.7) months. For the remaining 16% (21 patients), duration of PKDL was 16.6 (5.5) months, with over half of these cases showing either no change or worsening of PKDL during the first 12 months. Grade or severity of PKDL did not appear to influence the duration of PKDL and unless disease was very severe, these patients did not require treatment^34^. Although formal time to event data are not available, and other age groups have not been studied systematically, current clinical practice in Sudan is based on the premise that patients with persistent PKDL for six months or longer duration are not expected to rapidly self-resolve their lesions, and such patients are therefore provided with the standard of care, a protracted course of liposomal amphotericin B (AmBisome®; 2.5mg/kg/day for 20 days). Hence, as for other studies evaluating new drug regimens for PKDL^16,50,51^, patients with PKDL of greater than six months duration were enrolled for this study (median duration of PKDL of 30 months), with the expectation of a relatively stable disease over the short window of follow up. With the caveat that our present study contains no control arm for self-cure, we are encouraged by the finding that over a 3-4 month period approximately one third of patients resolved their PKDL lesions in the absence of further treatment, with a further 25% showing some clinical improvement. The extended and variable time frames over which clinical improvements in PKDL were observed was perhaps not surprising given the highly heterogeneous nature of PKDL in this study population. A randomised placebo-controlled trial (RCT) of ChAd63-KH in persistent PKDL patients in Sudan is currently underway (ClinicalTrials.gov Identifier: NCT03969134) to determine whether the clinical improvements observed in the current study are vaccine related. Whilst only ~20% of patients in Sudan present with persistent PKDL, in South Asia persistent disease reflects the norm and PKDL patients represent a significant risk to the regional VL elimination campaign^52-54^. The results from the current study also provide a clear incentive to evaluate therapeutic vaccination with ChAd63-KH as a tool for the management of PKDL cases in South Asia^55^.

Similar to healthy UK volunteers receiving the same vaccine, PKDL patients showed no unusual vaccine-induced responses, in keeping with their general state of health. Indeed, WBTA indicated that PKDL patients respond with a vigorous innate immune response, qualitatively similar to that seen in healthy UK volunteers^32^ in terms of GO term enrichment and predicted upstream regulators of gene expression. A direct quantitative comparison is not however possible due to the use of different platforms for WBTA analysis (Ion AmpliSeq™ Transcriptome Human Gene Expression Kit in this study vs. RNA-seq in ^32^). Module analysis suggested that adolescents with PKDL may generate more pronounced B cell responses to ChAd63-KH than adults, though the functional significance of this remains to be determined and the small sample size and variable demographics suggest this finding needs confirmation with a larger sample size and through more detailed phenotypic and functional analysis of the B cell response. Nevertheless, stronger humoral responses in adolescents have been observed previously with other vaccines e.g. the quadrivalent HPV vaccine^56^. Through a logistic regression model, we also identified a small number of modules with predictive power for identifying patients reaching the clinical endpoint of >90% improvement. Of note, module LI.M139 contains only 11 genes encoding endo/lysosomally-located proteins and enzymes, namely the scavenger receptor CD68, the catabolic enzyme acid glucosidase (GAA), cathepsins B, D, H and S (involved in various aspects of antigen presentation and myeloid cell activation^58^), proteins associated with cholesterol transport (NPC2) and glycolipid catabolism (PSAP), proteins involved in endo/lysosomal protein sorting (AP1S2 and SORT1) and the transporter SLC11A1 (a polymorphic divalent metal ion transporter associated with natural resistance to *Leishmania* and other intracellular pathogens and enhanced antigen presentation^59^). It remains to be determined to what extent this vaccine-induced molecular signature relates to vaccine efficacy or is reflective of underlying immunological or genetic differences in the response to vaccination in those patients destined to self-resolve their PKDL. The latter situation may be viewed as somewhat analogous to the association between leishmanin skin test reaction, self-resolution and/or response to therapy observed in PKDL patients^4,16,34^.

In conclusion, this study represents an important milestone in the development of a therapeutic vaccine as an additional tool for PKDL patient management, and more broadly encourages further exploration of therapeutic adenovirus-vectored vaccines for other infectious diseases.

## Data Availability

Transcriptomic data have been deposited in the Gene Expression Omnibus (GEO) repository with accession number GSE156645 and are available from: https://www.ncbi.nlm.nih.gov/geo/query/acc.cgi?acc=GSE156645.

## Acknowledgements

The clinical trial was funded by a Wellcome Trust Translation Award (WT108518MA; https://wellcome.ac.uk). Additional support for immunological studies was provided by a Wellcome Trust Senior Investigator Award (to PMK; WT104726) and by the TRANSVAC2 program supported by the European Union’s Horizon 2020 Research and Innovation programme under grant agreement No. 730964 (TNA1802-02; https://www.transvac.org). The funders played no role in study design or decision to publish.

## Author contributions

Conceptualization, AMM, CJNL, EAGK, PMK, RG, MB, TA Methodology, AMM, AML, BMY, CJNL, EAGK, FS, MO, PMK, SF and RW Investigation, AEAM, AK, AML, BMY, CJNL, EAGK, FS, MAAA, MO, LM, PMK and SF Writing – Original Draft, PMK

Writing – Review & Editing, AK, AMM, AML, CJNL, EAGK, MO, PMK, RG and TA Funding Acquisition, AMM, CJNL, EAGK, PMK, RG, MB, TA Resources, AMM, CJNL, EAGK, PMK Supervision, AMM, CJNL, EAGK, PMK

## Data availability

Supplementary Tables

**Table S1. Demographic, baseline and AEs summary for study participants.**

Sheet 1 provides demographic characteristics and biochemistry and haematology data at baseline. Sheet 2 summarises local and systemic AE number by cohort. Sheets 3 and 4 summarise AEs by relatedness to study intervention. Sheet 5 lists all reported AEs and grade.

**Table S2. Transcriptomics analysis raw data**

Sheet 1 contains count data for each patient at each time point examined. Sheet 2 contains patient descriptors and read depth and rate of target detection. Sheets 3 and 4 contain DE gene lists for adults (sheet 3) and adolescents (sheet 4) comparing those vaccinated with 7.5×1010 vp ChAd63-KH to pre-vaccination baseline.

**Table S3. Module descriptions and results of machine learning.**

Sheet 1 contains genes associated with each immune module used in this study.

Sheet 2 contains genes predicted to be associated with clinical response based on analysis using all modules or only modules DE at p>0.05.

Remaining sheets provide lists of the significantly enriched modules for each cohort at each time point and /or dose.

**Table S4. Gene set enrichment analysis**

Sheets contain output from EnrichR for GO Biological Processes 2018, Cellular Component 2018, Reactome 2016 and WikiPathways 2019 for DE genes at day 1 post vaccination in adult high dose and adolescent high dose cohorts.

